# Global prediction for monkeypox epidemic

**DOI:** 10.1101/2022.10.21.22280978

**Authors:** Li Zhang, Jianping Huang, Bin Chen, Yingjie Zhao, Danfen Wang, Wei Yan

## Abstract

The monkeypox epidemic has now spread all over the world and has become an epidemic of widespread concern in the international community. Before the emergence of targeted vaccines and specific drugs, it is necessary to numerically simulate and predict the epidemic. In order to better understand and grasp its transmission situation, and put forward some countermeasures accordingly, we predicted and simulated monkeypox transmission and vaccination scenarios using models developed for COVID-19 predictions. The results suggest the monkeypox epidemic will spread to almost all countries in the world by the end of 2022 based on modified SEIR model prediction. The total number of people infected with monkeypox will reach 100,000. The top five countries will be the United States, Brazil, Germany, France and Britain with more than 28000, 20000, 4000, 4500 and 4000 cases respectively. If 30% of the population is vaccinated, the number of infected people will drop by 35%.

## 1 Introduction

Monkeypox has infected more than 70,000 people worldwide and death cases are starting to appear by October 21, 2022. Furthermore, the monkeypox epidemic has been declared a global health emergency by the World Health Organization on May 23, 2022^1^. Monkeypox is a zoonotic infectious disease caused by the monkeypox virus in the genus Orthopoxvirus, which belongs to the same genus as smallpox virus. Typical symptoms include fever, severe headache, swollen lymph nodes, back pain, rash, muscle pain and so on^2^. Monkeypox can be transmitted to humans from rodents, primates and other animals, and also from humans. Monkeypox is spread by direct contact with the body fluids of infected people or with virus-contaminated items such as clothing and bedding. Similar to COVID-19, it can also be transmitted through highly toxic respiratory droplets. Studies have shown that the smallpox vaccine can prevent 85% of monkeypox virus^3^, but its acquisition has become more difficult because of the extinction of smallpox in 1980. In addition, there has been no breakthrough in the specific drug for monkeypox so far^4^. Therefore, due to the difficulties in development vaccines and specific drugs, non-pharmaceutical interventions are still an important means to deal with the monkeypox epidemic, similar to it of COVID-19. The mathematical models can provide a reliable scientific basis for various control measures. Moreover, in the context of increasingly severe climate change, the risk of zoonotic disease has increased by 58%^5^, and the development of general epidemic dynamic models is also an extremely important method to quickly respond to various potential large-scale infectious diseases in the future.

## 2 Method

To predict the global spread of monkeypox, we used the modified SEIR model (SEIQRDP) which divides the population into seven categories: susceptible people (S), potentially infected people (E, infected cases in a latent period), infected people (I, infected cases which have not been quarantined), quarantined people (Q, confirmed and quarantined cases), recovered people (R), deaths (D) and insusceptible people (people who take protective measures). The model consists of the following equations:

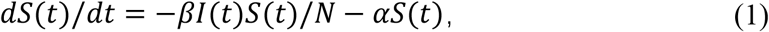

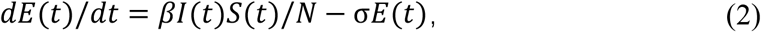

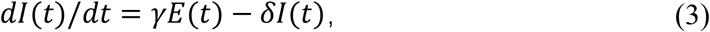

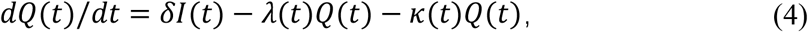

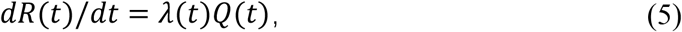

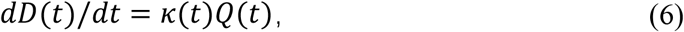

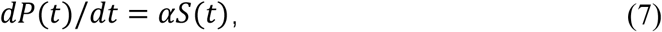

The total population (N) is equal to the sum of the seven populations at any time.

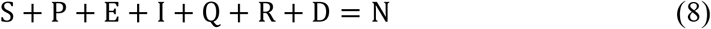

The coefficients of the model: α, β, γ, δ, λ, and κ represents the rate of protection, the rate of infection, the inverse of the average latent time, the rate at which infected people enter quarantine, the time-dependent recovery rate, and the time-dependent mortality rate, respectively. The model assumes that the total population is a constant, so the birth rate and death rate are not considered. Compared with the traditional SEIR model, this modified model adds P (insusceptible) and Q (quarantined) cases. In reality, people will take self-protection measures because they are aware of the existence of the epidemic, so the possibility of infection may be less. Moreover, the actual number of infected people is almost impossible to know, while the number of people in quarantine (official quarantine and self-quarantine) is. The number of death is also separated from the number of recoveries to obtain more accurate simulation and prediction result.

The data of monkeypox was obtained from the website: https://www.monkeypox.global.health/. It includes the data of all the countries with monkeypox outbreaks, and now there are more than 100 countries. By introducing the latest epidemic data in real time and using the improved least squares method to invert the latest model coefficients, the subsequent epidemic development can be predicted^6,7^.

## 3 Result

Figure 1a shows the cumulative predictions to the end of December 2022 for all countries with monkeypox epidemic so far. It can be seen from the figure that the monkeypox epidemic, like the COVID-19, has heterogeneity in spatial distribution. In North America, South America, and Europe, the outbreak is significantly more severe than in other parts of the world. Specifically, from figure 1b, the number of cases in the United States, Brazil, Germany, France, the United Kingdom and Italy will all exceed 1,000 by the end of December 2022. In particular, the United States and Brazil will have more than 20,000 cases. This is due to differences in culture, lifestyle, population density, control measures, etc. in different countries. Mass vaccination could help humans fight the monkeypox virus, if 20% of people are vaccinated against monkeypox by the end of the year, it could reduce cases by about 21%. And if the vaccination ratio is increased to 30%, the number of cases can be reduced by 35%. This suggests that the proportion of vaccinations and the proportion of potential case reductions are not simply linear. Although monkeypox is less contagious than COVID-19, it will still spread to many countries around the world due to a globalized transportation system. It is only a matter of time before most countries in the world will be invaded by monkeypox virus. Perhaps due to people’s experience in dealing with the COVID-19, people took corresponding protective measures after the large-scale outbreak of monkeypox. In addition, thanks to the development of the smallpox vaccine, people can be vaccinated against the monkeypox virus as a targeted manner. This also helps reduce potential monkeypox virus infection. These factors have led to a downward trend in the current monkeypox epidemic. However, this may only be the first wave of monkeypox outbreaks, and subsequent viruses may mutate, or there may be more waves as environmental conditions change. Regarding the current epidemic situation, it is safer to maintain a cautious attitude.

**Figure 1:**
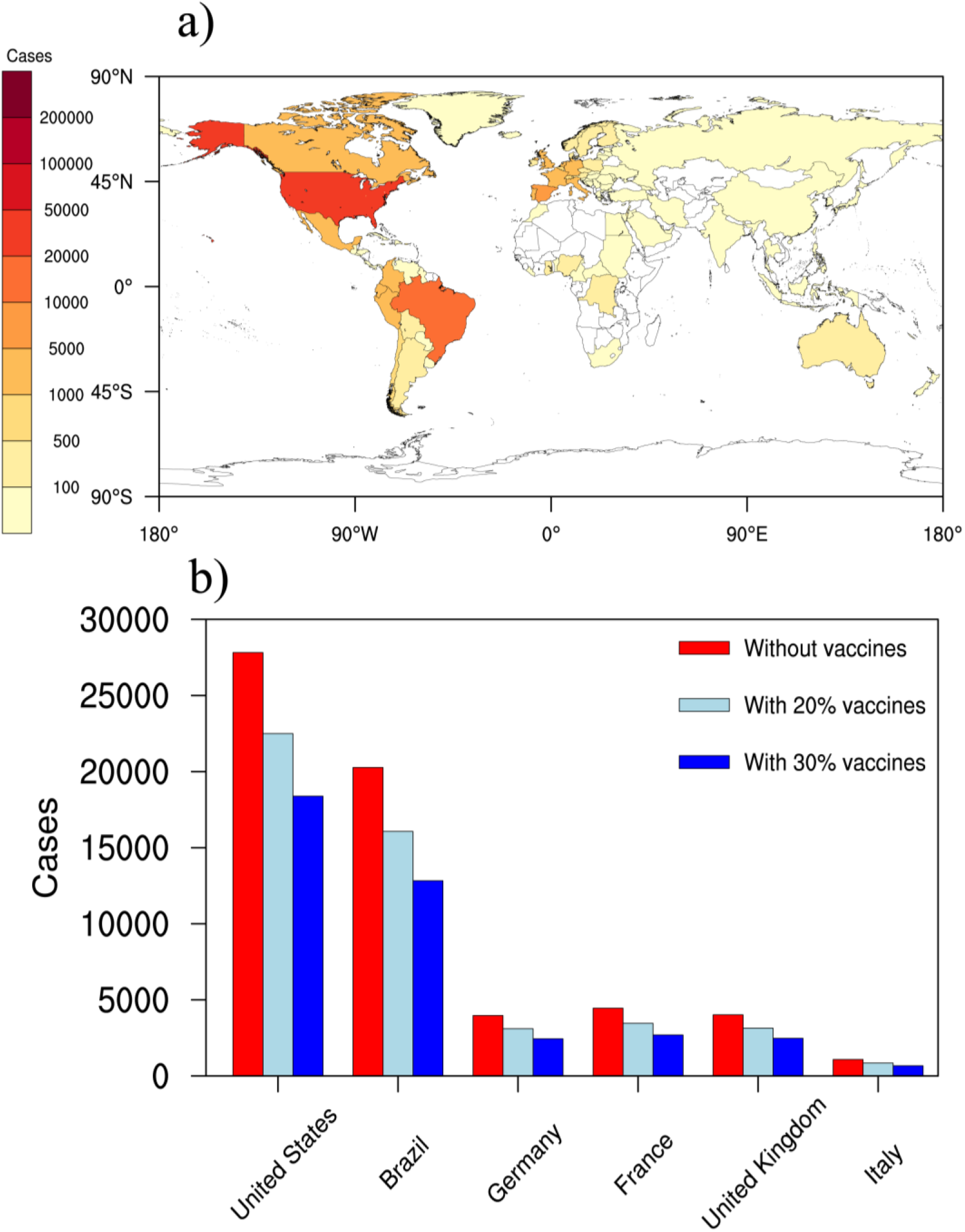
a, Global prediction of cumulative cases of Monkeypox on Dec 31, 2022; b, cumulative cases of Monkeypox on Dec 31, 2022 for six countries with or without vaccines.

## 4 Discussion

The prediction of monkeypox outbreaks can provide meaningful prediction and analysis on the global time and space scale, and can provide scientific basis for decision-making of government departments around the world. Although the current general model can make some basic predictions, there are still some aspects to be improved. Finally, our understanding of monkeypox virus is still relatively lacking, and further understanding of its transmission mechanism and mutation characteristics is needed. As climate change becomes more and more severe, the habitat of a large number of wildlife may change, potentially leading to more contact with humans, thus creating opportunities for a large number of pathogens to enter the human body. If humans are simply struggling to cope with a large-scale outbreak that has already occurred, and lack of monitoring research on pathogen spillover at the source, we may pay a very heavy price.

## Data Availability

All data produced in the present study are available upon reasonable request to the authors
All data produced in the present work are contained in the manuscript

https://www.monkeypox.global.health

